# Trends and spatial distribution of low-level viremia among Ugandan children and adolescents, 2014–2023

**DOI:** 10.64898/2025.12.17.25342004

**Authors:** Janet Kobusinge Lubega, Patrick Twesigye, Richard Migisha, Lilian Bulage, Benon Kwesiga, Proscovia Nambuya, Isaac Sewanyana, Eleanor Namusoke Magongo, Alex Riolexus Ario

## Abstract

**Background:** Low-level viremia (LLV) is as an early warning signal for treatment failure and onward transmission risk. Uganda has achieved major progress in expanding viral load (VL) testing coverage, yet the burden and trends of LLV among children and adolescents living with HIV (CALHIV) remain insufficiently described. This study assessed national LLV prevalence, temporal trends, and regional distribution from 2014–2023 to guide pediatric HIV program planning.

**Methods:** We analyzed routine VL data from Uganda’s Laboratory Information Management System (LIMS) housed at the Central Public Health Laboratories for CALHIV aged 0–19 years with VL results between 2014 and 2023. Viral suppression was defined as <200 copies/mL for plasma and <400 copies/mL for dried blood spot (DBS) samples. LLV was categorized as 201–999 copies/mL for plasma and 401–999 copies/mL for DBS. We analyzed LLV proportions over time and disaggregated them by demographic (age, sex), clinical (ART regimen, duration on ART, adherence, WHO stage), and facility-level (ownership, level of care) characteristics. The Mann-Kendall trend test was used to assess the significance of observed trends.

**Results:** A total of 974,873 VL tests were analyzed, of which LLV accounted for 14% (138,126). Nationally, LLV increased from 10.5% in 2014 to 16.2% in 2023 (p<0.001). LLV rose in both males (10.7%→16.9%, p<0.001) and females (10.3%→15.6%, p=0.025), with the steepest increase among infants (12.7%→22.5%, p<0.001). Increases were observed among CALHIV on first-line regimens (10.2%→15.7%, p<0.01), those on ART ≥5 years (12.5%→17.8%, p<0.01), and those with good adherence (10.2%→14.9%, p<0.001). In 2023, five of fifteen regions recorded LLV prevalence above 20%.

**Conclusion:** The rising LLV burden among CALHIV in Uganda signals emerging virologic challenges that may compromise long-term treatment outcomes. Strengthening VL monitoring, early detection of LLV, and tailored adherence interventions are critical to sustaining viral suppression and preventing progression to high-level viremia.

## Background

Despite significant investments in pediatric Antiretroviral Therapy (ART) programs and early infant diagnosis, only 46% of children on ART have achieved viral suppression, far below the adult rates of suppression and the global target of 95% suppression (1, 2).

They face higher risks of loss to follow-up, poor adherence, and delayed disclosure of HIV status, all of which hinder treatment outcomes (3). This persistent gap highlights critical weaknesses in pediatric HIV care and underscores the urgent need for targeted interventions to improve ART adherence, access to optimized regimens, and routine viral load (VL) monitoring among children.

In low- and middle-income countries, nearly 20% of people living with HIV experience LLV and 9% remain virologically non- suppressed. Children and adolescents are disproportionately affected, with prevalence rates higher than those observed among adults (4). Low level viremia is consistently associated with an increased risk of non-virologic suppression across all populations (5). For children and adolescents, this is particularly concerning as neither group has achieved viral suppression levels above 95% despite improvements in ART coverage over the years (6). Low level viremia represents an intermediate state between full suppression and high-level viremia and serves as an important early warning marker of impending virological failure. Monitoring LLV therefore provides a critical opportunity for timely action before a child or adolescent progresses to virological failure. These challenges underscore the need for age-responsive programming that incorporates enhanced VL monitoring and prompt interventions whenever elevated VLs are detected including detectable LLV (7).

In Uganda, challenges along the pediatric HIV care cascade persist. In 2022, of the estimated 95,000 children under 15 living with HIV, only 60% achieved viral suppression (1). Earlier national data from the 2020 Uganda Population-based HIV Impact Assessment (UPHIA) reported even lower suppression rates of 32.3% among children aged 0–14 years and 49.4% among adolescents aged 15–19 years highlighting the substantial gaps that remain (8). Although a national VL monitoring program has been in place since 2014, evidence on LLV in this vulnerable population remains limited. Studies in other sub-Saharan African countries, including Tanzania and South Africa, have shown LLV to be strongly predictive of treatment failure (9, 10). However, Ugandan evidence is still scarce. Addressing this knowledge gap is critical for guiding age-responsive programming and timely clinical intervention.

Following the 2013 WHO guidelines, many countries in the region, including Uganda, define viral non-suppression as having ≥1,000 HIV RNA copies/mL, with VL testing conducted at six months after ART initiation, at 12 months, and annually thereafter for suppressed adults, while suppressed children and adolescents are monitored every six months (11). Non-suppressed children and adolescents living with HIV(CALHIV) follow a well-defined clinical pathway involving intensive adherence counselling (IAC) and potential regimen switch, LLV patients remain on current regimens, with limited clinical action beyond counseling (12). In 2023, WHO explicitly classified LLV, defined as 200–999 copies/mL, as detectable but still suppressed under the programmatic threshold of ≤1,000 copies/mL. Therefore, LLV does not meet the formal definition of non-suppression, even though it represents a detectable incomplete viral control and carries potential risks of drug resistance and progression to treatment failure (7). With the rising LLV prevalence and evidence of their poor outcomes progression, this minimal-intervention approach has come under increasing examination (13). Without targeted action, opportunities to intervene before full treatment failure may be missed, making LLV surveillance a critical frontier in pediatric and adolescent HIV care. This study utilized national Laboratory Information Management System (LIMS) data from 2014 to 2023 to assess LLV among children and adolescents aged 0–19 years living with HIV in Uganda. We described the trends and spatial distribution of LLV among CALHIV, Uganda, 2014–2023 to inform HIV programming.

## Methods

### Study design, setting, and data source

We conducted a descriptive analysis of routinely collected VL monitoring data for CALHIV in Uganda between January 2014 and December 2023 obtained from the National Laboratory Information Management System (LIMS) database managed by the Central Public Health Laboratories (CPHL).

Uganda is an East African country with 45.9 million people, over half aged <18 years, and a national HIV prevalence of 5.1% (14). HIV care and treatment services for CALHIV are provided free of charge through a decentralized health system. Health facilities range from Health Center (HC) IIs to national referral hospitals, alongside private-not-for-profit and accredited private facilities (15). HC IIs (parish level) provide outpatient HIV services including HIV testing and counselling and receive outreach support from higher HCs for CALHIV. HC IIIs (sub-county level) provide outpatient HIV services including HIV testing and counselling, ART initiation and refills for clinically stable clients, and routine follow-up including VL monitoring. HC IVs (county level) and hospitals provide comprehensive outpatient and limited inpatient HIV care, while district, regional, and national referral hospitals provide specialized care for complex patients (15). In addition, decentralized adolescent and pediatric HIV clinics are available at most of the HCs, offering designated spaces or clinic days for CALHIV aged 0–19 years and these clinics are staffed by trained clinicians, expert clients, and peer counsellors to provide tailored services for this population (16).

The dataset used for this analysis was accessed on 9 August 2024 and was extracted in de-identified form. Only system-generated LIMS codes were included, which were not linked to names or other personal identifiers and could not be used to identify individuals. Authors did not have access to any direct personal identifying information during or after data collection.

### National Viral Load Monitoring Program, 2014–2023

The national VL monitoring program, introduced in 2014, is coordinated by the Central Public Health Laboratories (CPHL) through a hub-and-spoke sample transport system linking peripheral health facilities to regional and national laboratories (17). Clinicians at ART health facilities request VL tests using the standardized Health Management Information System (HMIS) laboratory request form. For each VL test, the form captures demographic information including age and sex and clinical information including ART regimen, duration on ART, tuberculosis status, and pregnancy or breastfeeding status. It also records WHO clinical stage and adherence score, both of which reflect the clinician’s judgment at the time of patient review.

Indications for VL testing are guided by national policy and therefore categorized according to the timing VL is requested. For adults, VL testing is routinely recommended at 12 months after ART initiation. This is categorized as a routine VL test at 12 months and annually thereafter for those who are stable, categorized as routine monitoring. In contrast, CALHIV undergo VL monitoring every six months because of their higher risk of treatment failure and adherence challenges (12, 18) categorized as a routine VL test at 6 months. Additional VL requests are made at the first antenatal care (ANC) visit to monitor maternal suppression and prevent mother-to-child transmission (12) categorized as a routine VL test at ANC. Non-routine VL tests are performed for clients who are not stable. Non-routine VL tests are performed for clients who are not stable and are categorized as suspected failure. These also include VL testing after intensive adherence counselling (IAC), categorized as VL after IAC, to help distinguish adherence-related non-suppression from true treatment failure. Clinicians may also request repeat VL testing at their discretion for closer follow-up of unstable clients, categorized as repeat VL. A clinician can also suspect failure and provides a reason for failure on the request form. When a VL test is requested because of suspected treatment failure, the reason for the request is further categorized as clinical failure (persistent HIV-related symptoms or opportunistic infections), immunological failure (declining CD4 counts or poor immune recovery), or virological failure (persistent elevated VL despite good adherence). Routine monitoring performed also categorized and documented as a reason for VL testing.

Blood samples for VL testing are collected at ART health facilities across Uganda, using both dried blood spot (DBS) and plasma. Samples are transported through the hub-and-spoke network to laboratories, and results are uploaded into LIMS to support surveillance, program monitoring, and clinical decision-making (19).

While national VL monitoring began in 2014, formal recognition of LLV was only introduced in 2020, when Uganda updated its guidelines. Before this, reporting focused only on suppression (<1000 copies/mL) and non-suppression (≥1,000 copies/mL)(16). From 2020 onward, LLV, defined as 201–999 copies/mL for plasma and 401–999 copies/mL for DBS, was introduced as a distinct reporting category to identify CALHIV at risk of virological failure (19, 20).

### Study variables, data abstraction

We obtained individual-level viral-load data for CALHIV from the LIMS, for the period 2014–2023. Each VL record included both the laboratory result, routinely collected demographic, and clinical information. From the records, all VL results were categorized into three outcomes: suppressed (<200 copies/mL for plasma or <400 copies/mL for DBS), low-level viremia, and high-level viremia (HLV, ≥1,000 copies/mL) to determine their proportions in relation to LLV. For this analysis, only LLV (201–999 copies/mL for plasma; 401–999 copies/mL for DBS) was retained as the primary outcome. After categorizing VL outcomes and identifying LLV as the primary outcome, we applied additional data cleaning procedures to address potential duplication and missing information. Because children are expected to have at least two VL tests annually, multiple records per individual were common. However, patient identifiers were not consistently available or standardized in LIMS, limiting our ability to reliably determine whether repeat tests belonged to the same child. As a result, all LLV records were retained in the analysis. Records with missing key information (age, sex, or VL result) and biologically implausible values were excluded during data cleaning.

Other study variables included were demographic, clinical, and programmatic characteristics of CALHIV. Demographic variables were age, sex, and geographic region. Age was analyzed as a continuous variable and also grouped into developmental categories (0–1, 2–5, 5–9, 10–14, and 15–19 years). Geographic region was classified using Uganda’s 15 programmatic regions.

Clinical variables included were pregnancy/breastfeeding status, ART regimen line (first-line, second-line, third-line), duration on ART (<6 months, 6–11 months, 1–2 years, 2–5 years, ≥5 years), WHO clinical stage (I–IV), adherence (good, fair, poor), tuberculosis (TB) status (yes/no), and TB treatment phase (initiation or continuation).

Testing indication was categorized as routine monitoring (routine, six months after ART initiation, 12 months after ART initiation, or first antenatal care visit) or targeted monitoring (suspected treatment failure, repeat after IAC, or other reasons). In patients with suspected treatment failure were categorized as clinical, immunological, or virological failure. Programmatic variables were specimen type (plasma or DBS), health facility level, and facility ownership (public or private).

### Data analysis

Data were analyzed using Stata/SE version 14.0 (StataCorp LP, College Station, TX, USA). All eligible VL records were exported from LIMS and cleaned by removing biologically implausible results and excluding records missing key variables (age, sex, or VL result). Categorical variables were summarized as frequencies and percentages, while continuous variables such as age were described using medians and interquartile ranges (IQRs). Low level viremia was calculated as the proportion of valid VL results that fell within the LLV range. The variable for reason for failure was divided onto two categorizes: routine and non-routine. Non-routine included for analysis were clinical, immunological, or virological failure. Temporal trends in LLV were demonstrated using line graphs and assessed for monotonic (increasing or decreasing) trends, using the Mann–Kendall trend test, with analyses stratified by age group, sex, and geographic region. Spatial distribution of LLV was assessed using Quantum Geographic Information System (QGIS) software, version 3.8.2-Zanzibar (QGIS Development Team, 2019, Open Source Geospatial Foundation Project to generate regional maps. Statistical significance was defined at p<0.05.

## Results

### Proportion of low-level viremia among children and adolescents, Uganda, 2014–2023

From 2014 and 2023, a total of 974,872 VL test results from CALHIV were analyzed. Of these, 67% (650,471) were suppressed, 14% (138,160) had LLV, and 19% (186,241) had HLV.

### Distribution of low-level viremia by demographic characteristics of children and adolescents, Uganda, 2014–2023

Among the 138,160 CALHIV with LLV, the median age was 11 years (IQR: 7–15). Most of the CALHIV with LLV were aged 10–14 years 35% (48,256) or 5–9 years 28% (38,925), and over half were female 55% (73,821). Nearly all CALHIV with LLV were from government facilities 98% (n=134,964) and by level of care, more than half were from HCIIIs 52% (71,307Among female adolescents with LLV, less than 1% were pregnant 0.6% (788) or breastfeeding 0.9% (1,196) at the time of VL testing (Table 1).

**Table 1:**
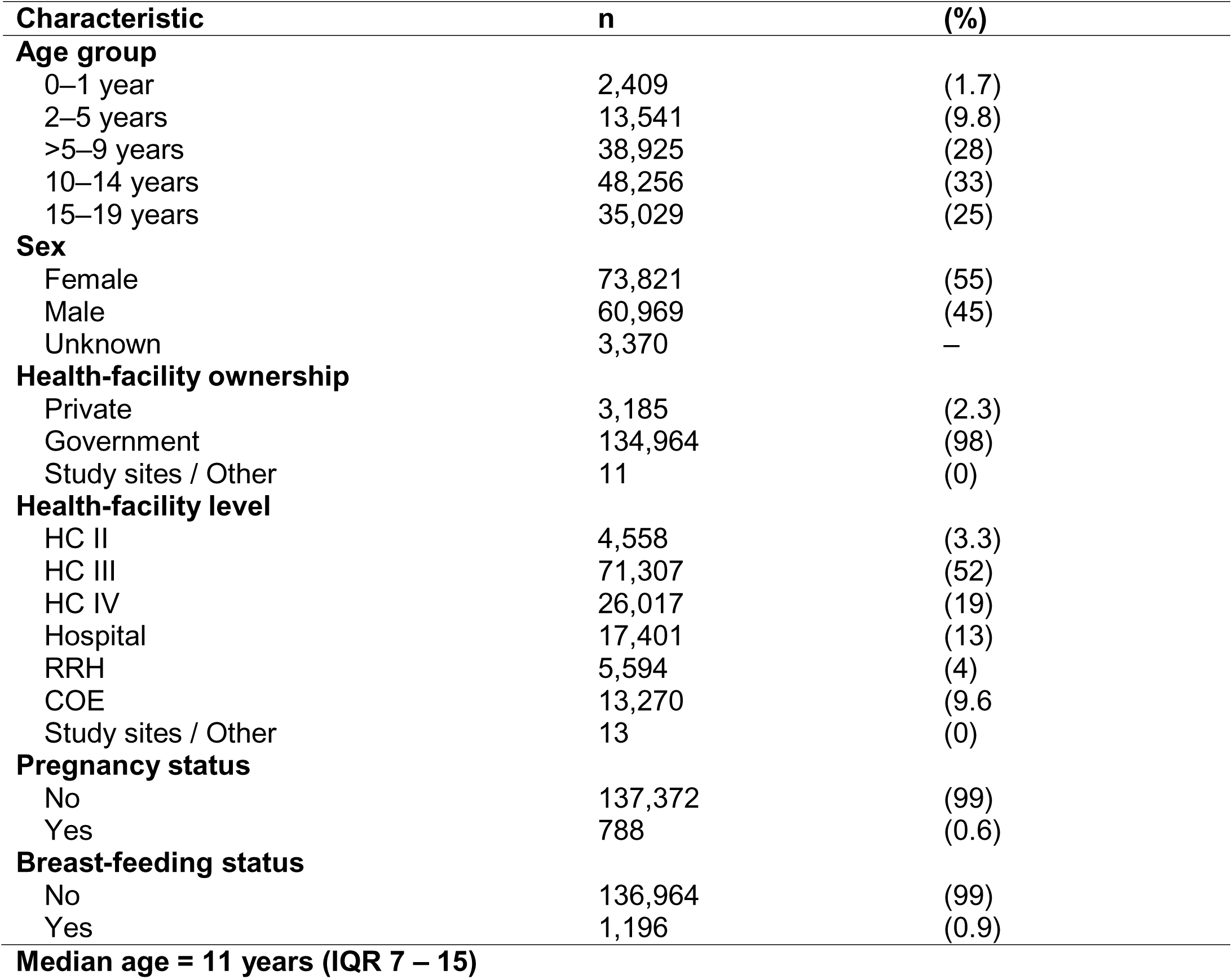
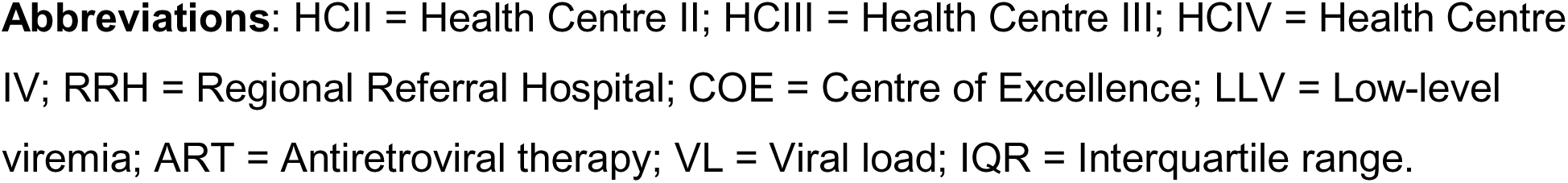
Distribution of low-level viremia by demographic characteristics of children and adolescents, Uganda, 2014–2023 (N = 138,160)

### Distribution of low-level viremia by clinical characteristics of children and adolescents, Uganda, 2014–2023

Among CALHIV with LLV, the majority were on first-line ART 87% (112,259), had good adherence 93% (127,377), and were in WHO Stage I 95% (42,832). Nearly half had been on ART ≥5 years 42% (54,597). Most CALHIV with LLV were identified through routine monitoring 89% (123,476 while for suspected failure (clinical, immunological, and virological) only accounted for <5%. (Table 2).

**Table 2:**
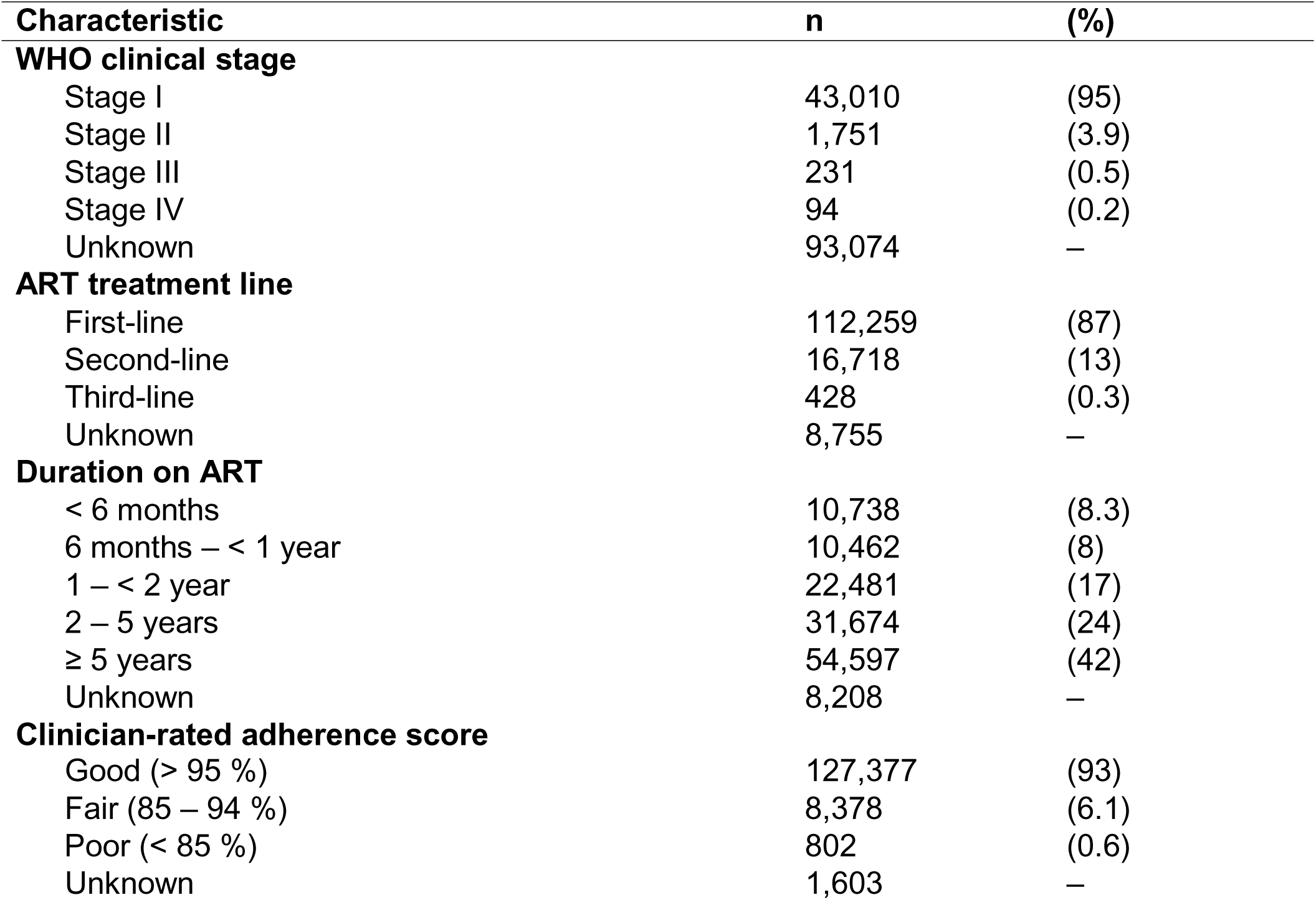
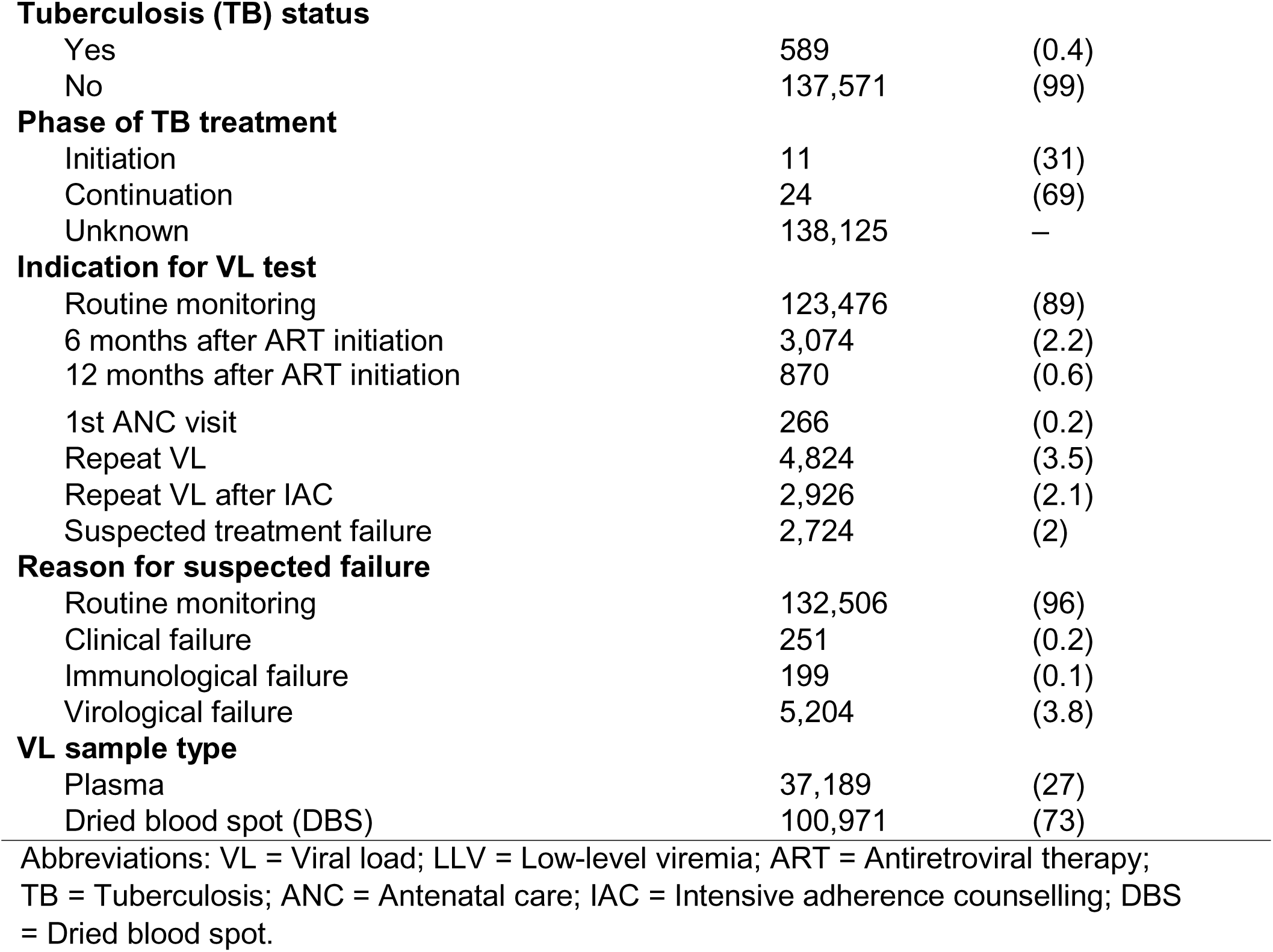
Distribution of low-level viremia by clinical characteristics of children and adolescents, Uganda, 2014–2023 (N = 138,160)

| **Characteristic** | **n** | **(%)** |
| --- | --- | --- |
| **WHO clinical stage** |  |  |
| Stage I | 43,010 | (95) |
| Stage II | 1,751 | (3.9) |
| Stage III | 231 | (0.5) |
| Stage IV | 94 | (0.2) |
| Unknown | 93,074 | – |
| **ART treatment line** |  |  |
| First-line | 112,259 | (87) |
| Second-line | 16,718 | (13) |
| Third-line | 428 | (0.3) |
| Unknown | 8,755 | – |
| **Duration on ART** |  |  |
| < 6 months | 10,738 | (8.3) |
| 6 months – < 1 year | 10,462 | (8) |
| 1 – < 2 year | 22,481 | (17) |
| 2 – 5 years | 31,674 | (24) |
| $\geq 5$ years | 54,597 | (42) |
| Unknown | 8,208 | – |
| **Clinician-rated adherence score** |  |  |
| Good (> 95 %) | 127,377 | (93) |
| Fair (85 – 94 %) | 8,378 | (6.1) |
| Poor (< 85 %) | 802 | (0.6) |
| Unknown | 1,603 | – |
| **Tuberculosis (TB) status** |  |  |
| Yes | 589 | (0.4) |
| No | 137,571 | (99) |
| **Phase of TB treatment** |  |  |
| Initiation | 11 | (31) |
| Continuation | 24 | (69) |
| Unknown | 138,125 | – |
| **Indication for VL test** |  |  |
| Routine monitoring | 123,476 | (89) |
| 6 months after ART initiation | 3,074 | (2.2) |
| 12 months after ART initiation | 870 | (0.6) |
| 1st ANC visit | 266 | (0.2) |
| Repeat VL | 4,824 | (3.5) |
| Repeat VL after IAC | 2,926 | (2.1) |
| Suspected treatment failure | 2,724 | (2) |
| **Reason for suspected failure** |  |  |
| Routine monitoring | 132,506 | (96) |
| Clinical failure | 251 | (0.2) |
| Immunological failure | 199 | (0.1) |
| Virological failure | 5,204 | (3.8) |
| **VL sample type** |  |  |
| Plasma | 37,189 | (27) |
| Dried blood spot (DBS) | 100,971 | (73) |

Abbreviations: VL = Viral load; LLV = Low-level viremia; ART = Antiretroviral therapy; TB = Tuberculosis; ANC = Antenatal care; IAC = Intensive adherence counselling; DBS = Dried blood spot.

### Trends in low level viremia among children and adolescents, Uganda, 2014–2023: Overall patterns and disaggregation by sex, age group, and health facility level

From 2014 and 2023, the overall proportion of LLV among children and adolescents aged 0–19 years rose by 26%, with three phases of increase: from 0% to 11% between 2014 and 2018, 15% in 2019, and 12% to 26% between 2021 and 2023 (p<0.001) (Figure 1A). Low level viremia declined among children aged 2–4 years, from 16% in 2014 to 10% in 2023 (p<0.001). In contrast, LLV increased among adolescents aged 10–14 years, from 24% to 36% over the same period (p<0.001) (Figure 1B). LLV remained stable among females at 53% in both 2014 and 2023 (p=0.12) and declined slightly among males from 47% to 44% (p=0.72), with neither change statistically significant (Figure 1C). LLV declined at Centers of Excellence from 85% in 2014 to 4% in 2023 (p<0.0001) and at regional referral hospitals from 17% to 2% (p<0.001). In contrast, HCIIIs showed a significant increase, from 14% in 2015 to 62% in 2023 (p<0.0001) (Figure 1D).

**Figure 1:**
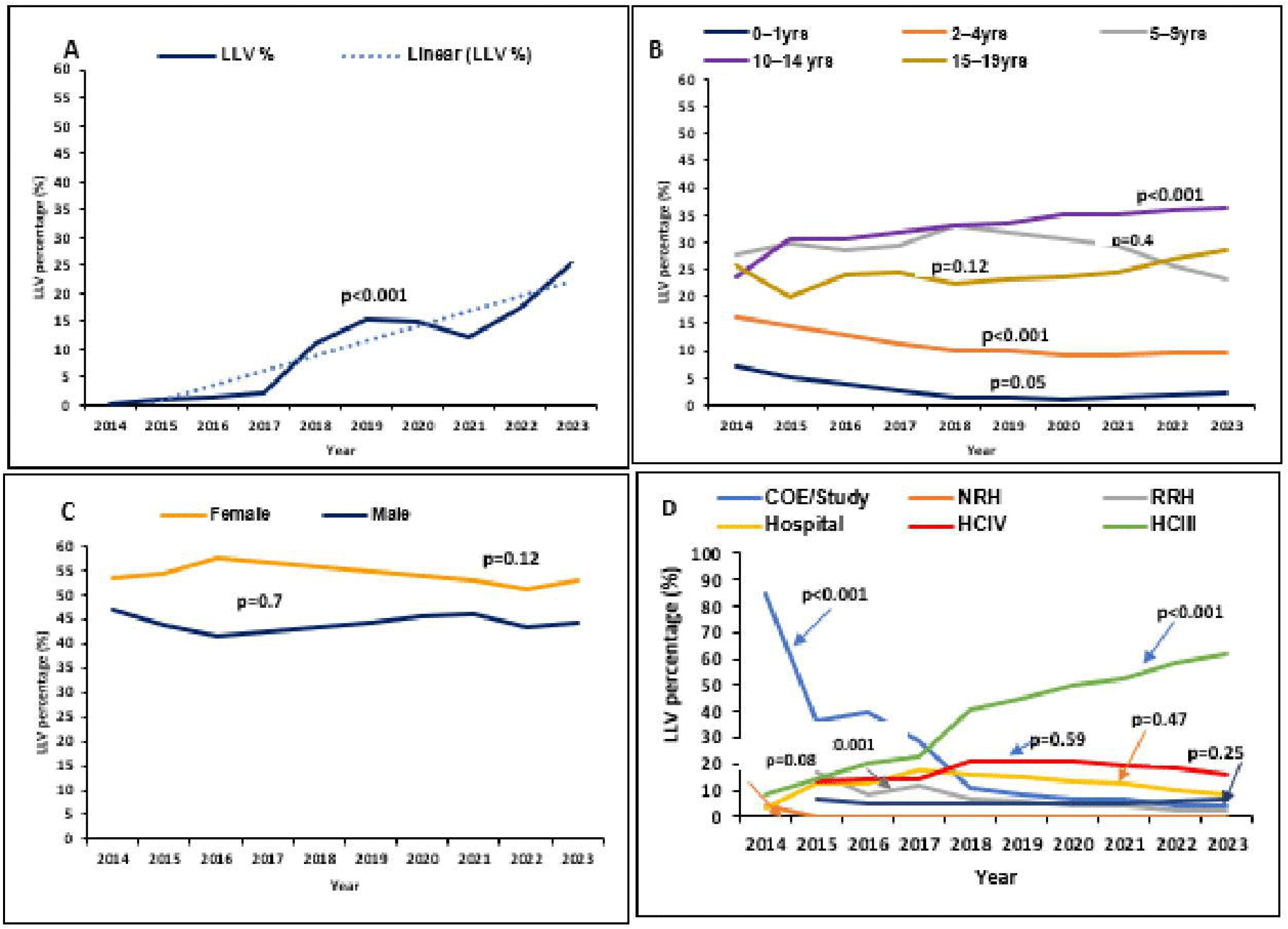
Trends of low-level viremia among children and adolescents aged 0–19 years, Uganda, 2014–2023. Overall trends (A), and disaggregated by age group (B), sex (C), and (D) health facility level

### Trends in low-level viremia among children and adolescents aged 0–19, Uganda, 2014-2023, disaggregated by treatment line, duration on antiretroviral therapy, adherence level, and World Health Organization clinical stage

Between 2014 and 2023, LLV on first-line ART rose from 80% to 91% but showed no significant trend (p=1). In second-line users, LLV declined from 30% to 0% (p=0.37), while in third-line users it remained consistently below 5% (p=0.32) (Figure 2A).

**Figure 2:**
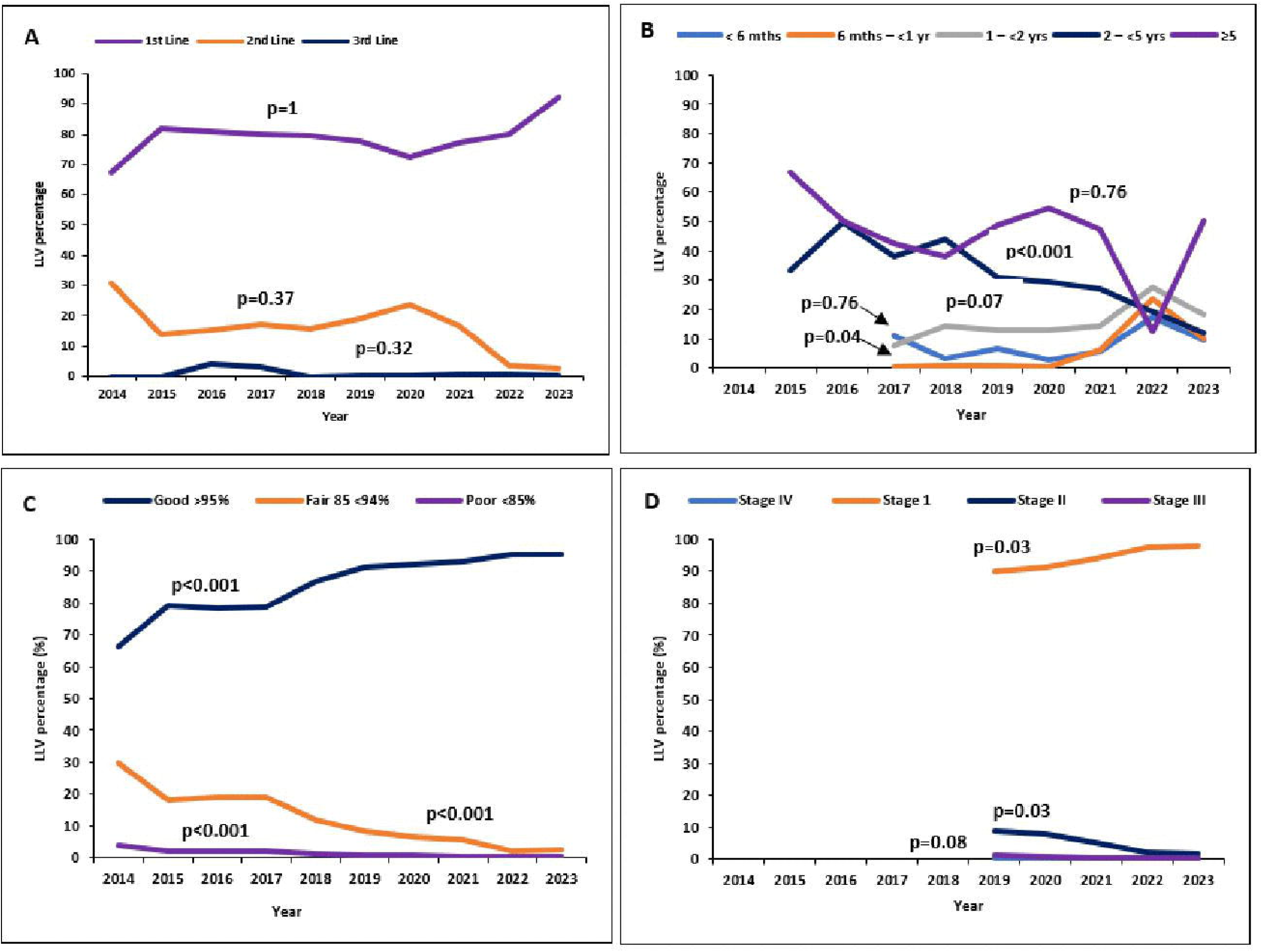
Trends in low-level viremia among children and Adolescents, Uganda, by treatment Line (A), duration on ART (B), ARV adherence Level (C), and WHO clinical stage (D), 2014–2023

Low-level viremia increased among those on ART for 6 months–<1 year (p=0.03) and showed a borderline increase for 1–<2 years (p=0.07). A significant decline was observed among those on ART for 2–<5 years (p<0.001) (Figure 2B).

Among those with good adherence (>95%), LLV rose from 66% in 2014 to 95% in 2023 (p<0.001). It declined among those with fair adherence (85–94%), from 30% to 3% (p<0.001), and among those with poor adherence (<85%), from 4% to 0% (p<0.001) (Figure 2C). From 2019 to 2023, LLV among CALHIV categorized as Stage I increased from 90% to 98% (p=0.03), while those in Stage II declined from 9% to 2% (p=0.03) (Figure 2D).

### Spatial distribution of low-level viremia among children and adolescents, Uganda, 2014–2023

Between 2014 and 2023, the spatial distribution of LLV in Uganda showed marked regional variation. The sharpest increases occurred in Tooro (0% in 2014 to 14% in 2023), North Central (0% in 2014 to 11% in 2023), and South Central (2% in 2014 to 16% in 2023), with North Central reaching 13–14% between 2018 and 2020 and South-Central peaking at 17% in 2017. South Central and North Central consistently recorded the highest LLV levels after 2016, while Karamoja remained lowest, rarely exceeding 1%, with Bukedi and Bugisu also maintaining <5% throughout. Mid-range trends were seen in Ankole, Bunyoro, Busoga, and Acholi, all gradually increasing to 6–11% in later years. (Figure 3)

**Figure 3.**
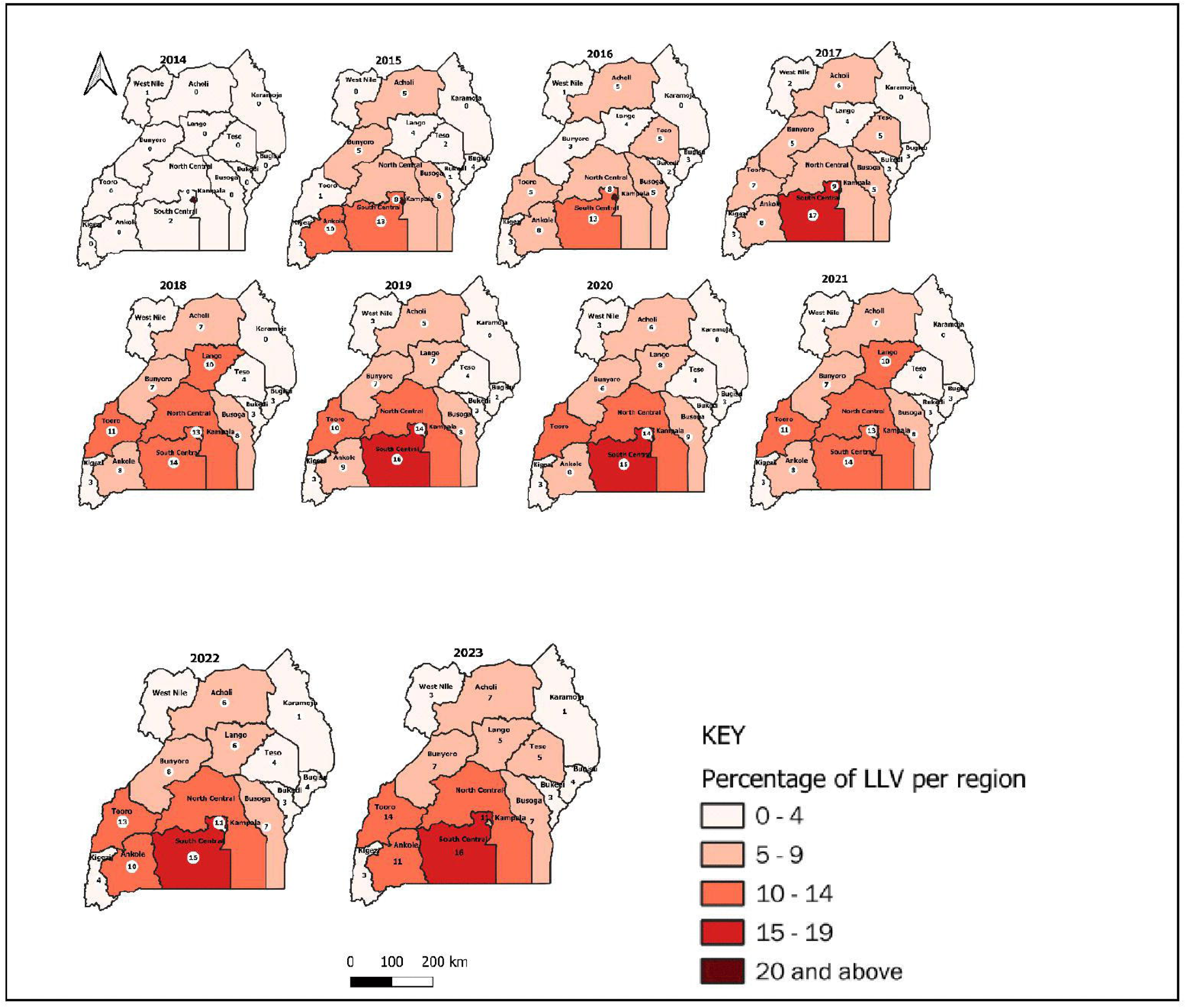
: Spatial trends in low-level viremia among children and adolescents, Uganda, by region, 2014–2023

## Discussion

In this national descriptive analysis of viral load data from 2014–2023, we found that LLV among Ugandan children and adolescents living with HIV (CALHIV) accounted for ∼14% of results and increased by ∼26% over the decade. Low-level viremia was concentrated in adolescents aged 10–14 years, with minimal sex differences. Most LLV occurred on first-line ART, was highest among long-term ART users (≥5 years), and increased during early treatment (6–12 months). Despite being recorded as having “good” adherence and WHO Stage I at the time of testing, many still had LLV.

Low-level viremia was reported mainly from government facilities especially Health Centre IIIs and was geographically concentrated in South Central and North Central, with consistently low levels in Karamoja. These patterns of LLV signal a need for rapid repeat VL, more objective adherence assessment, and earlier regimen review to protect long-term suppression.

From 2014 and 2023, the overall proportion of LLV among CALHIV increased steadily, showing distinct phases of rise that reflected evolving program implementation and monitoring over the decade. These overall trends indicate that rising LLV coincided with the scale-up of routine VL testing and decentralization of ART services, alongside formal recognition of LLV in national and WHO guidance respectively (16, 17, 19, 20). The overall increase in LLV among CALHIV mirrors patterns seen in Ugandan adult cohorts, where LLV rose from 2016 to 2020 and was strongly predictive of subsequent virologic non-suppression (13). Similar increases have also been reported in other sub-Saharan African settings (9, 10). These findings reinforce the growing recognition of LLV as a programmatic concern and align with calls from studies in South Africa and Kenya to reconsider the >1,000 copies/mL threshold for defining virologic failure (22).

The consistency of these trends across diverse populations highlights LLV as an early warning indicator that should not be overlooked in pediatric and adolescent care.

Age-specific patterns showed LLV was predominantly observed among in adolescents aged 10–14 years, while younger age groups declined consistent with adolescence-specific barriers such as incomplete disclosure, stigma, and competing priorities, which undermine adherence (23, 24). Younger children, by contrast, benefit from more intensive caregiver involvement and structured follow-up. Similar age-associated LLV patterns have been reported in other pediatric cohorts in sub-Saharan Africa (25), while Ugandan adult studies demonstrate LLV clustering in younger adults (26), suggesting that LLV vulnerability shifts with age. These findings emphasize adolescents as a key group for intensified adherence and psychosocial support interventions.

Variation in LLV was evident across treatment line and duration. Low-level viremia was predominant among first-line ART, and declined among second-line users, consistent with the higher resistance barrier of boosted PI regimens and closer monitoring after switches (27). Adult data from Uganda confirm that LLV predicts subsequent virologic failure (13, 26), underscoring the importance of addressing LLV early through enhanced adherence support and timely regimen review in line with national and WHO guidelines (12, 20). These results indicate that first-line ART remains the programmatic “pressure point” for pediatric LLV and should be prioritized for early intervention. Early LLV (6–12 months after initiation) increased, and long-term ART (≥5 years) carried the greatest burden patterns consistent with risks from delayed initial suppression, adherence fatigue, and archived resistance (25, 28, 29). These findings suggest that both early and long-term ART users require tailored programmatic responses to prevent persistence of LLV. Programmatically, LLV detected on first-line should trigger rapid repeat VL and timely regimen review in line with national/WHO guidance.

Most LLV occurred among clients recorded as having “good” adherence, possibly highlighting limitations of routine adherence measures (self-report, pill counts) and “white-coat” effects that can overestimate true adherence (30, 31, 32, 33).

Low-level-viremia was most frequent in WHO Stage I, emphasizing that apparent wellness does not exclude viraemia. In the ART era, VL-based monitoring remains the preferred standard. These findings highlight that LLV represents a programmatic signal of hidden adherence gaps, reinforcing the need for objective adherence tools even among children recorded as adherent. Clinically, LLV was predominantly observed among children who appeared well, while WHO Stages II–IV were uncommon. This underscores the limited utility of clinical staging in detecting viraemia in the ART era(34). Low-level viremia in clinically stable children emphasizes the critical role of routine VL monitoring and the need to act on detectable viremia even in asymptomatic children (25). Programmatic reliance on clinical staging alone may therefore miss early signs of virologic failure in children.

Health facility level and geography influenced LLV detection. Low-level viremia was concentrated in government facilities and Health Centre IIIs, consistent with decentralization and expanded access (7, 17, 36). Regionally, South Central and North Central had the highest LLV burden, while Karamoja remained lowest patterns that likely reflect differences in underlying HIV burden, service capacity, and laboratory networks (8, 15, 19, 20, 36). Targeted support for adolescents and long-term ART users at decentralized public sites in high-burden regions, alongside strengthened follow-up after routine VL milestones, is essential to sustain suppression and advance 95-95-95 goals

## Study limitations

The major limitations of this study were use of routinely collected national VL data, where incomplete or missing records may have affected the results, and inability to link repeat tests at the individual level. Children and adolescents are expected to have at least two viral load tests per year, and repeat tests are often done when results are delayed; however, because patient identifiers were not consistently available or standardized in LIMS, we could not reliably determine whether repeat tests belonged to the same individual. Consequently, some children with multiple tests may have been counted more than once, and we could not determine whether LLV was a temporary finding that later suppressed (transient) or a sustained pattern over multiple tests (persistent) so a longitudinal study was not possible. This also meant we could not follow viral load changes over time at the individual level. In addition, the cross-sectional design does not allow for assessment of causality. Despite these limitations, the study used one of the largest pediatric and adolescent HIV viral load datasets, providing important insights for similar low-resource settings.

## Conclusion

The rising LLV burden among CALHIV in Uganda signals emerging virologic challenges that may compromise long-term treatment outcomes. Strengthening VL monitoring, early detection of LLV, and tailored adherence interventions are critical to sustaining viral suppression and preventing progression to high-level viremia. Sustained investment in decentralized viral load testing, adherence support, and timely regimen review will be essential to maintain viral suppression gains and advance Uganda’s 95-95-95 targets.

## List of abbreviations

ACP: AIDS Control Program
AIDS: Acquired Immunodeficiency Syndrome
ANC: Antenatal Care
AHD: Advanced HIV Disease
ART: Antiretroviral Therapy
ARV: Antiretroviral
CALHIV: Children and Adolescents Living with HIV
CDC: U.S. Centers for Disease Control and Prevention
COE: Center of Excellence
CPHL: Central Public Health Laboratories
DBS: Dried Blood Spot
DHIS2: District Health Information Software version 2
EMTCT: Elimination of Mother-to-Child Transmission
HIV: Human Immunodeficiency Virus
HLV: High-Level Viremia
HMIS: Health Management Information System
IAC: Intensive Adherence Counselling
HC II: Health Centre II
HC III: Health Centre III
HC IV: Health Centre IV
LIMS: Laboratory Information Management System
LLV: Low-Level Viremia
NRH: National Referral Hospital
PEPFAR: U.S. President’s Emergency Plan for AIDS Relief
PMTCT: Prevention of Mother-to-Child Transmission
QGIS: Quantum Geographic Information System
RNA: Ribonucleic Acid
UNAIDS: Joint United Nations Programme on HIV/AIDS
UNFPA: United Nations Population Fund
UNICEF: United Nations Children’s Fund
UPHIA: Uganda Population-based HIV Impact Assessment
US: United States
VL: Viral Load
WHO: World Health Organization

## Ethical considerations

We used retrospective, routinely collected surveillance data, and no direct engagement with participants occurred. The Uganda Public Health Fellowship Program, Uganda National Institute of Public Health, Ministry of Health, Kampala, Uganda, as part of the National Rapid Response Team, is mandated to access and analyze national surveillance and laboratory data, including DHIS2 and other routine public health datasets, to inform outbreak response and public health programming. We were granted permission for access to and analysis of these de-identified data and authorized dissemination of findings through scientific publications by the Ministry of Health, Kampala, Uganda, We abstracted data that was de-identified prior to analysis, stored on password-protected computers, and accessed only by members of the investigation team.

The U.S. Centers for Disease Control and Prevention (CDC), Atlanta, Georgia, USA, provided a Non-Research Determination (NRD), indicating that this activity does not constitute human subjects research and that its primary intent was to improve public health practice and disease control. This determination was made in accordance with the International Guidelines for Ethical Review of Epidemiological Studies by the Council for International Organizations of Medical Sciences (1991) and the Office of the Associate Director for Science, CDC. The activity was reviewed by CDC and conducted consistent with applicable federal law and CDC policy (e.g., 45 C.F.R. part 46; 21 C.F.R. part 56; 42 U.S.C. § 241(d); 5 U.S.C. § 552a; 44 U.S.C. § 3501 et seq.). The study was performed in accordance with the Declaration of Helsinki as it applies to public health surveillance activities.

## Consent for publication

Not applicable.

## Data availability

The datasets on which our findings are based belong to the Uganda Public Health Fellowship Program, MoH and CPHL. For confidentiality reasons, the datasets are not publicly available. However, the datasets are available upon reasonable request from the corresponding author with permission from the Uganda Public Health Fellowship Program and MoH. Requests can be directed at.

## Funding and disclaimer

The project was supported by the President’s Emergency Plan for AIDS Relief (PEPFAR) through the United States Centers for Disease Control and Prevention Cooperative Agreement number GH001353-01 through the Makerere University School of Public Health to the Uganda Public Health Fellowship Program, Ministry of Health.

The findings and conclusions in this report are those of the authors and do not necessarily represent the official views of the US Centers for Disease Control and Prevention, the Department of Health and Human Services, the Agency for Toxic Substances and Disease Registry, the Makerere University School of Public Health, or the Uganda Ministry of Health.

## Authors’ contribution

JKL conceptualized the idea, analyzed and interpreted the data, and drafted the manuscript. EM supervised and along with RM, BK, LB, PM, IS, and ARA reviewed the manuscript for intellectual content.

## Acknowledgements

We thank the staff of the Uganda Public Health Fellowship Program for the technical support and guidance offered during this study. We would like to thank staff of the AIDS control Program (Ministry of Health) for the technical guidance given during the conceptualization stage.

